# The effect of the definition of ‘pandemic’ on quantitative assessments of infectious disease outbreak risk

**DOI:** 10.1101/2020.10.02.20205682

**Authors:** Benjamin J Singer, Robin N Thompson, Michael B Bonsall

## Abstract

In the early stages of an outbreak, the term ‘pandemic’ can be used to communicate about infectious disease risk, particularly by those who wish to encourage a large-scale public health response. However, the term lacks a widely accepted quantitative definition. We show that, under alternative quantitative definitions of ‘pandemic’, an epidemiological metapopulation model produces different estimates of the probability of a pandemic. Critically, we show that using different definitions alters the projected effects on the pandemic risk of key parameters such as inter-regional travel rates, degree of pre-existing immunity, and heterogeneity of transmission rates between regions. Our analysis provides a foundation for understanding the scientific importance of precise language when discussing pandemic risk, illustrating how alternate definitions affect the conclusions of modelling studies. This serves to highlight that those working on pandemic preparedness must remain alert to the variability in the use of the term ‘pandemic’, and provide specific quantitative definitions when undertaking one of the types of analysis that we show to be sensitive to the pandemic definition.

## 1 Introduction

In the early stages of an infectious disease outbreak, it is important to determine whether the pathogen responsible may go on to cause an epidemic or a pandemic [1–5]. There is extensive literature on determining the probability of a major epidemic given a small population of initial infected hosts [6–9]. This research has produced a natural mathematical definition of an epidemic, based on the bimodal distribution of outbreak sizes given by simple epidemiological models when *R*_0_ is larger but not close to one [10]. The term ‘pandemic’ has no corresponding theoretical definition, and there is no consensus mathematical approach to determining the probability of a pandemic. In this study, we examine how alternative definitions of ‘pandemic’ affect quantitative estimates of pandemic risk assessed early in an infectious disease outbreak.

The term ‘pandemic’ is used extensively, appearing in phrases such as ‘pandemic preparedness’ [11–13], ‘pandemic influenza’ [14–16], and ‘pandemic potential’ [17–19]. A Google Scholar search returns 25,800 results using the term ‘pandemic’ for 2019 alone.

The International Epidemiology Association’s Dictionary of Epidemiology defines a pandemic as “an epidemic occurring worldwide, or over a very wide area, crossing international boundaries and usually affecting a large number of people” [20]. Notably this definition makes an explicit reference to national borders. Contrastingly, a World Health Organisation (WHO) source makes reference to a pandemic as “the worldwide spread of a new disease.” [21] The use of the word ‘new’ here is ambiguous in the context of infectious diseases. HIV/AIDS is often referred to as a global pandemic, but is certainly not new on the time scale of, say, the emergence of influenza strains [22, 23]. A study by Morens et al. in 2009 finds that there is little in common between all disease outbreaks that have been referred to as pandemics, except that they have a wide geographical extension [24].

These kinds of differences between pandemic definitions can often go unnoticed, but in certain circumstances they can cause confusion between different stakeholders (e.g. between scientists and governments, or governments and the public), who may not have a shared background understanding of the term. In 2009, the World Health Organisation (WHO) declared a pandemic of H1N1 influenza, using criteria related to the incidence and spread of the virus in different WHO regions [25]. The criteria did not include reference to morbidity or mortality [26]. This fact led to some controversy over whether the declaration of a pandemic was appropriate, as it led some governments to mount an intensive response to an outbreak that resulted in fewer deaths than a typical strain of seasonal flu [27–30].

International health organisations such as the WHO have not provided any formal definitions of the term ‘pandemic’, and the WHO no longer uses it as an official status of any outbreak [25, 31]. It would, however, be hasty to dismiss the importance of the term on this ground. Although the WHO no longer uses the term ‘pandemic’ officially, the WHO Director-General drew attention to their use of the term as recently as March 2020, to describe the status of the COVID-19 outbreak [32]. The Director-General cited “alarming levels of inaction” as one of the reasons to use the term, along with the caveat that “describing the situation as a pandemic does not change WHO’s assessment of the threat posed by this virus”. The WHO’s use of the term was of interest to the public, receiving extensive press coverage [33–35]. The term ‘pandemic’ clearly continues to be important to indicate serious risk during disease outbreaks.

Regardless of the extent to which the pandemic definitions currently in use do or do not agree, they are all qualitative in nature, using descriptions such as “very wide area” and “large number of people”. Perhaps as a result of this, many quantitative studies on pandemics do not make use of a quantitative definition of a pandemic, but instead focus on causally related concepts, such as sustained transmission [19], or emergence of novel viruses [36]. Others treat the spread of a pathogen at a pandemic level as a context in which to study transmission dynamics, without paying special attention to how those dynamics lead to a pandemic as distinct from an epidemic or a more limited outbreak [37–39]. In this paper, we examine the effects of alternate pandemic definitions on the analysis of key epidemiological questions. The results provide a foundation for deciding the appropriate quantitative definition of ‘pandemic’ in a given context.

We make use of a metapopulation model to investigate the effects of pandemic definition on the results of a quantitative assessment of the probability of a pandemic. Metapopulation models are commonly applied to pathogens that spread between regions of the world, and so are appropriate for modelling pandemics [40–45]. We explore two different kinds of pandemic definition, following Morens et al. 2009 [24], specifically:

- the family of *transregional* definitions, where a pandemic is defined as an outbreak in which the number of regions experiencing epidemics meets or exceeds some threshold number *n*. We refer to specific transregional definitions as *n-region* transregional definitions, e.g. a 3-region transregional definition.
- the *interregional* definition, where a pandemic is defined as an outbreak in which two or more non-adjacent regions experience epidemics.

Note that these definitions require a specific sense of ‘region’. These regions could be countries, or they could be larger or smaller than individual countries—from counties to health zones to WHO regions. Our metapopulation model (detailed in the Methods section below) can be used to model regions of any size. We have chosen not to include definitions with criteria relating to the number of people infected or killed, instead of, or in addition to, geographical extension. Extension is the only universal factor in pandemic definitions, and so is the focus of the current study [24].

Three questions that feed into public health policy at the beginning of an outbreak are:

- Would interventions restricting travel reduce the risk of a pandemic?
- Does a portion of the population have pre-existing immunity, and does this affect pandemic risk?
- How is the risk of a pandemic affected by regional differences in transmission?

Using our metapopulation model, we explore how changing the pandemic definition does or does not affect our answers to these questions. We show that the precise definition of a pandemic used in modelling studies can (but does not always) affect the inferred risk. The predicted effects of travel restrictions, the influence of pre-existing immunity, and the impact of regional differences in transmission can all vary when alternative definitions of ‘pandemic’ are used. This demonstrates clearly the need to consider carefully the pandemic definition used to assess the risk from an invading pathogen. This is necessary for clear communication of public health risk.

## 2 Results

### 2.1 Travel rates

One important question about pandemic risk is what effect inter-regional travel rates have on the probability of a pandemic occurring [16, 17, 46, 47]. Here we model epidemics occurring in regions connected on a network whose connections and weighting can be set at fixed values representing the rates of travel between regions. We choose the simplest networks that can illustrate the effects of our different pandemic definitions—namely, the star network, in which one central region is connected to all others with equal weighting and the non-central regions lack any other connections, and the fully connected network, in which each region is connected to every other with equal weighting. Figure 1 illustrates that the connectivity of the full network is much higher than that of the star network. Using the star network allows us to make the distinction between adjacent and non-adjacent regions, thus allowing us to distinguish between transregional and interregional pandemic definitions.

**Figure 1:**
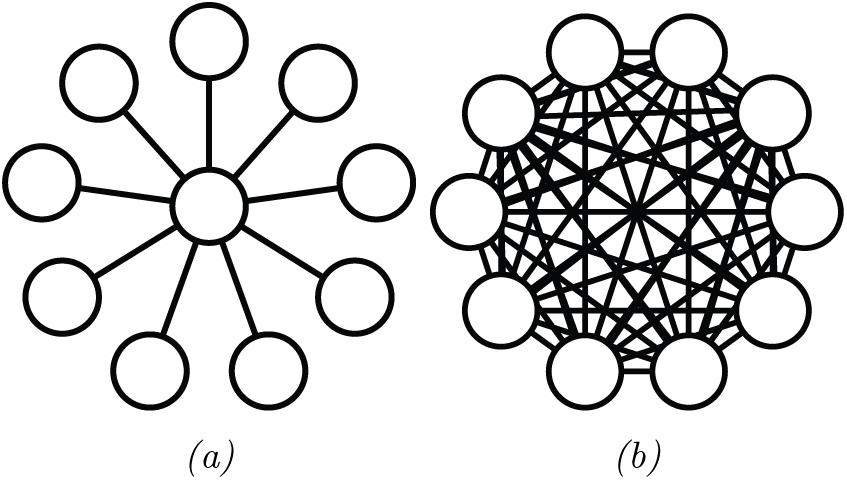
Illustrations of a) a star network and b) a full network, each with ten regions. Circles represent regions, and straight lines represent travel routes between regions.

Unless otherwise stated, all figures in the current study are generated with a transmission rate of *β* = 0.28 per day, a recovery rate of *µ* = 0.14 per day, and an inter-regional travel rate of 2 *×* 10^−4^ per day. This corresponds to a basic reproduction number (*R*_0_) of 2. These values are within the plausible range for both seasonal and pandemic influenza, and as such they plausibly simulate a pathogen of pandemic potential [38]. We further assume an initial population of 1000 susceptible individuals in each region, and that the disease is seeded by a single infectious individual in one arbitrary region.

Using a model with ten regions allows us to test a range of different transregional definitions of a pandemic. The pandemic probability under a range of *n*-region transregional definitions for a 10-region network with a variety of travel rates are shown in figure 2. An *n*-region transregional definition effectively provides a threshold number *n*—if more than *n* regions experience epidemics, the outbreak is counted as a pandemic, and otherwise it is not. Thus we indicate the different possible *n*-region definitions through their threshold numbers in figures 2, 5, and 6.

**Figure 2:**
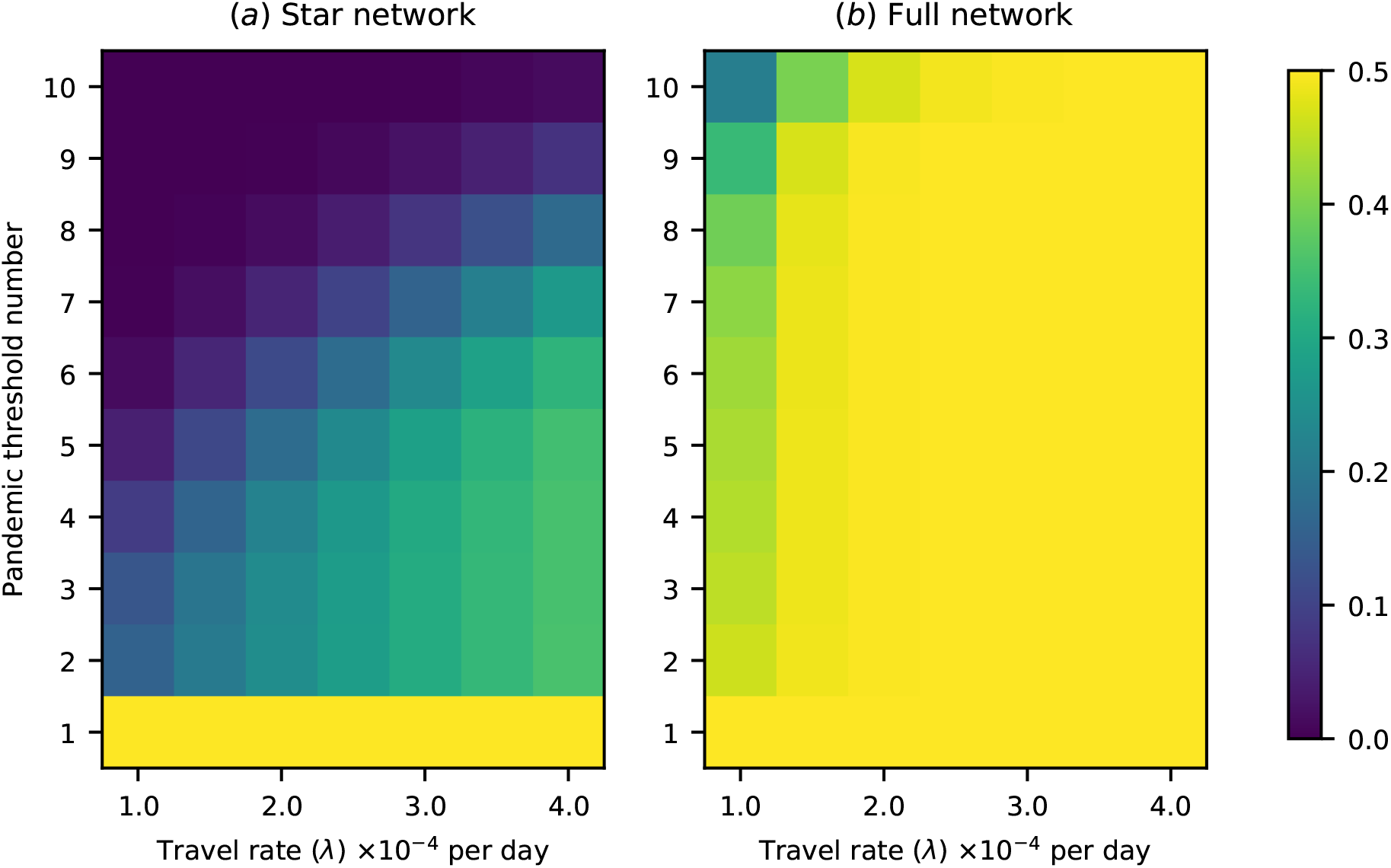
Pandemic probability for a range of between-region transmission rates and a range of pandemic definitions in terms of number of regions experiencing epidemics. The “pandemic threshold number” refers to the minimum number of regions that must experience epidemics before a pandemic is declared. Pandemic probability is, in general, sensitive to the pandemic definition used, but the degree of sensitivity depends on network structure and travel rates. a) Pandemic probability for a star network. Pandemic probability is, in general, highly sensitive to the pandemic definition used. b) Pandemic probability for a fully connected network. The sensitivity of pandemic probability to the pandemic definition used is significantly reduced at high travel rates.

The 1-region transregional definition merges the definitions of ‘pandemic’ and ‘epidemic’ in an implausible way, but it is included in these figures for comparison. The comparison between the probability of pandemics according to the 2-region definition and according to the 10-region definition shows the difference between pandemic definitions that are satisfied by any transregional transmission and definitions that are satisfied only by truly global spread. For the star network, or for the fully connected network with low travel rates, there is a marked difference between the probability of either of these definitions being satisfied. However, for the fully connected network at medium or high rates of travel, the probability of a pandemic reaches the maximum of 0.5 (i.e. 1 − 1*/R*_0_) and the definition makes little difference to the calculated risk. For any definition, the probability of a pandemic increases with the connectivity of the network, and with travel rates across the network.

We can also look at the difference in pandemic probability between the transregional and interregional definitions, which make use of a distinction between adjacent and non-adjacent regions. This is shown for a 10-region star network in figure 3a. We choose a star network as it is one of the simplest network types in which there are adjacent and non-adjacent regions. There is a difference between the two definitions, but the difference is much smaller than that between the interregional and 10-region (global) definition, and reduces as travel rates increase. In the case of a fully connected network, all regions are essentially adjacent to each other, so we compare only binary transregional and global definitions. We find that the definitions are highly distinct for low travel rates, but as this increases the difference between the likelihood of a pathogen causing an epidemic in one region and the likelihood of it causing epidemics in all regions disappears. This is due to the fact that the pathogen can be introduced into any population from any other.

**Figure 3:**
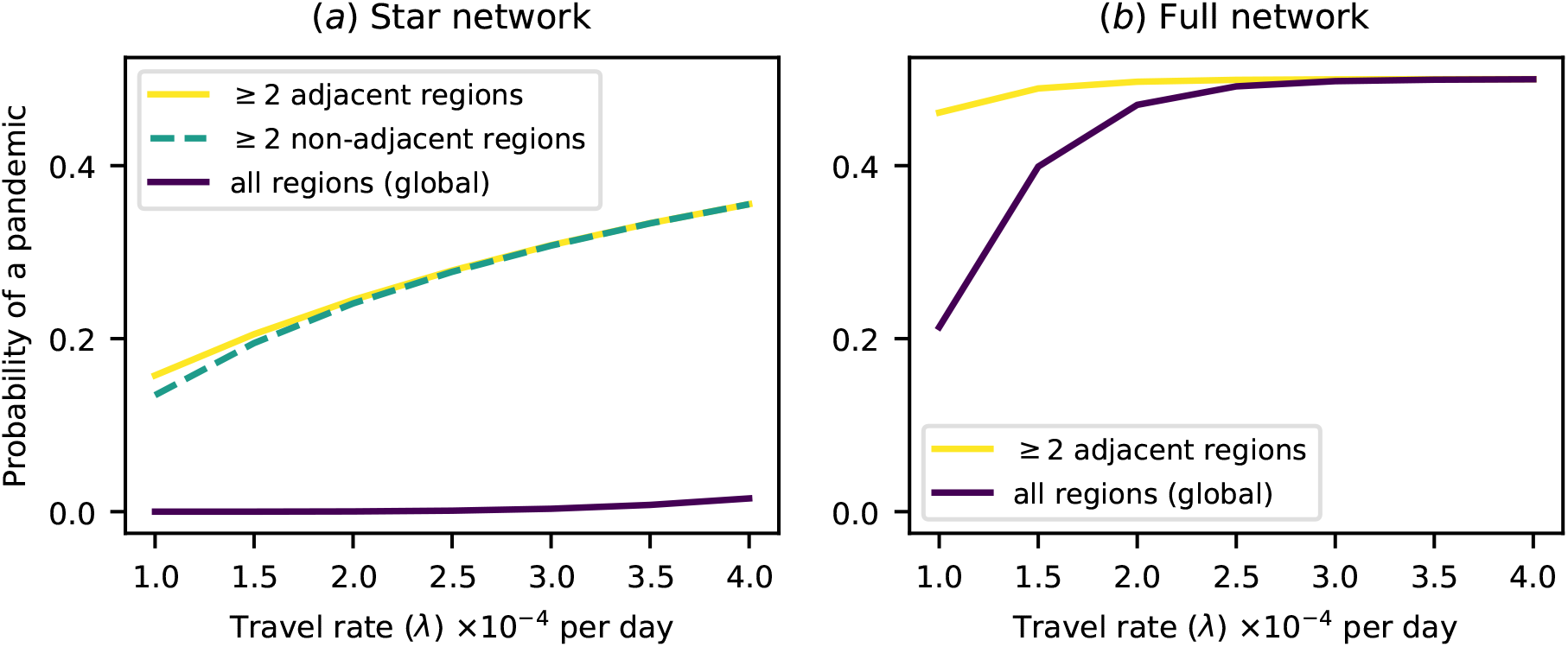
Plots of pandemic probability against travel rate for a range of pandemic definitions. The difference in probability for different pandemic definitions changes as travel rates increase. a) Plot of pandemic probability for a star network. b) Plot of pandemic probability for a full network. For a fully connected network all regions are adjacent, so no line is shown for non-adjacent regions.

In this section we have shown that, when a pandemic is defined in terms of which regions experience epidemics of a disease, different definitions can produce very different estimates of pandemic probability at low connectivity or travel rates, but have a much smaller effect at high connectivity and travel rates. In general, adjacency requirements make little difference to these results.

### 2.2 Cross-immunity

Some pathogens with pandemic potential have a prior history of infecting humans, such as pandemic influenza. Newly emerged pathogens with no history of infecting humans are less likely than these established pathogens to encounter regions where susceptible individuals have partial immunity to infection. Established pathogens may encounter individuals with partial immunity acquired from infections with previously circulating strains—i.e. *cross-immunity* [48, 49]. It can be important in responding to an outbreak to consider whether any individuals might have existing immunity. We can therefore investigate the interaction between immunology and pandemic definition by examining how cross-immunity affects our calculation of pandemic probability on a network.

We modelled the spread of a disease over a six-patch network with no cross-immunity initially, where the initial infected individual could appear in any region. We only included cases where at least one region experienced an epidemic of this initial pathogen. To simulate the emergence of a strain with higher pandemic potential, we then introduced a second pathogen with a higher transmission rate of *β* = 0.42 (corresponding to a basic reproduction number of 3), to which infection with the initial pathogen conferred some degree of partial immunity to infection *α*. See the Methods section for details of how our modelling framework handles cross-immunity. We defined a pandemic as occurring when all six regions experienced epidemics of the second pathogen, and repeated the model for two values of cross-immunity at a variety of between-region travel rates. The results are presented in figure 4.

**Figure 4:**
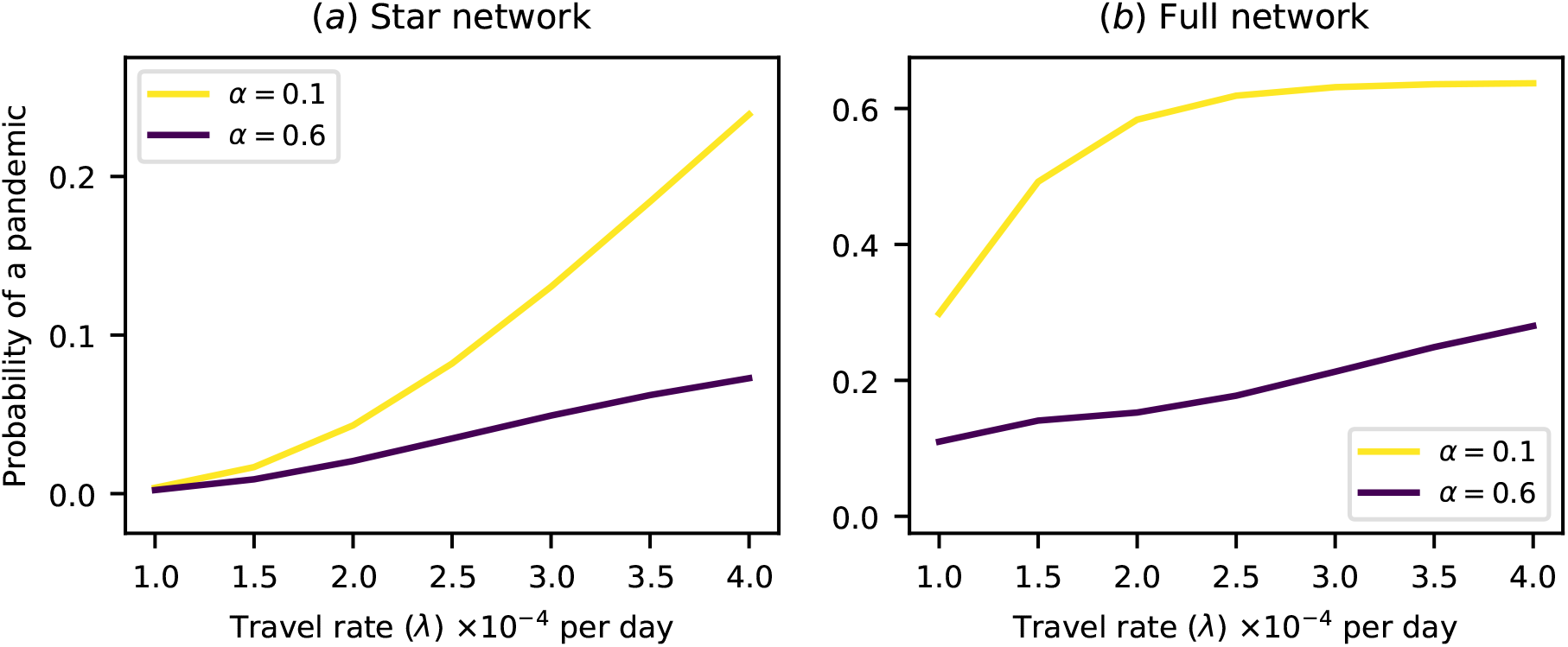
Plots of pandemic probability against travel rates for high and low values of cross-immunity (*α*) on six-region networks. A pandemic is defined here as all six regions experiencing epidemics. The plots show a large relative difference both in likelihood of pandemics and in how that likelihood scales with travel rates. The initial infected individual for each outbreak appears in a randomly chosen region. a) Plot of pandemic probability for a star network. b) Plot of pandemic probability for a full network.

First, increasing cross-immunity decreases the probability of a pandemic. Second, the presence of cross-immunity changes how pandemic probability scales with travel rates. In general, the probability grows faster with travel when the cross-immunity is low, except when it reaches a point of saturation as in figure 4b.

Figure 5 shows the simultaneous effects of different *n*-region transregional pandemic definitions and the degree of cross-immunity in determining the pandemic probability. Here we fix the travel rate at 2.0 *×* 10^−4^ per day. In the full network there is sudden transition from higher risk to lower risk, as cross-immunity approaches one. However, in the star network there is less circulation of the initial pathogen, so the effect of cross-immunity is less dramatic. Increased cross-immunity can also increase the difference in risk for different pandemic definitions—for the fully connected network, once cross-immunity reaches around *α* = 0.6, differences in probability between different thresholds become visible that are much smaller at lower values. This suggests that the declaration of a pandemic may be more sensitive to the exact pandemic definition for outbreaks of pathogens that encounter pre-existing immunity than for pathogens which encounter only fully susceptible populations. However, this effect is not seen for the star network, in which the low connectivity of the network results in larger differences in probability between different thresholds even at low cross-immunity.

**Figure 5:**
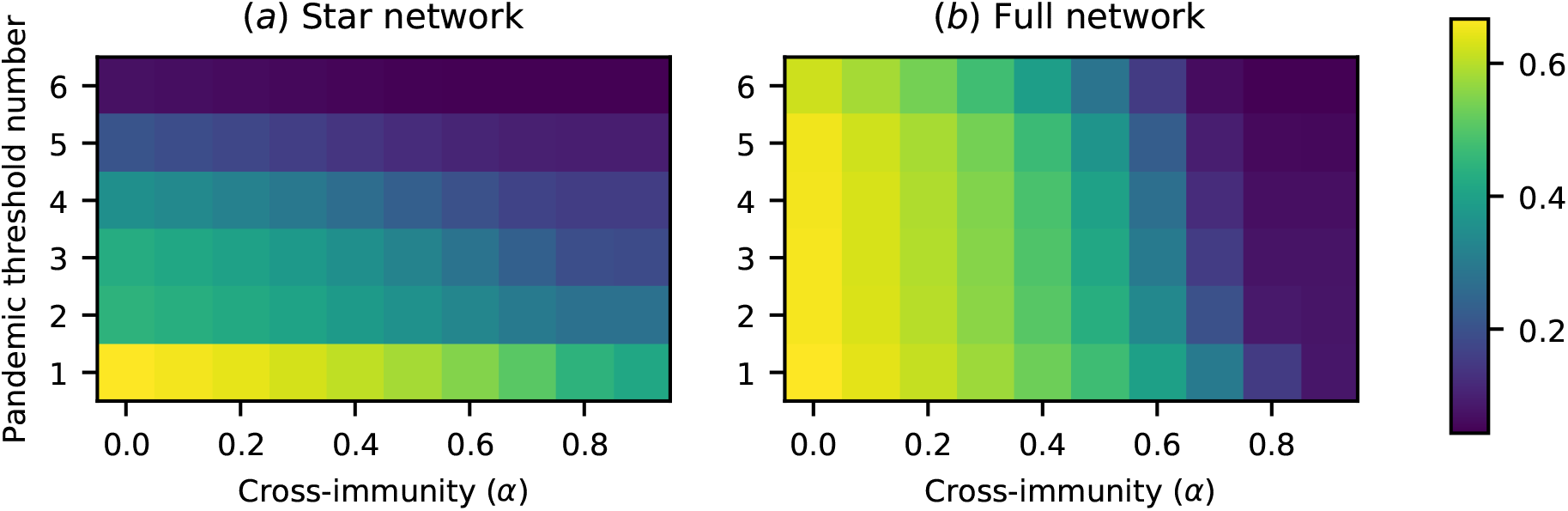
Pandemic probability for various degrees of cross-immunity (*α*) and a range of pandemic definitions in terms of number of regions experiencing epidemics, on a six-region network. a) Pandemic probability for a star network. b) Pandemic probability for a fully connected network. Here the sensitivity of pandemic probability to the pandemic definition used increases with cross-immunity, until the probabilty of any epidemic becomes very low.

### 2.3 Heterogeneous transmission

A topic of great concern during a pandemic is heterogeneity in risk between different countries or regions [50, 51]. Cross-immunity can create one kind of heterogeneity, since it is common for previous exposure to a pathogen to differ between regions. Another kind of heterogeneity is that due to different public health interventions. Here we ignore cross-immunity and instead examine a heterogeneous fully connected network of six regions, three of which have a higher rate of transmission of the pathogen than the other three. This can be thought of as an approximation to the difference between poor regions with a relative lack of public health interventions, and wealthy regions with well-funded public health organisations.

The level of heterogeneity was defined as the ratio of the transmission rate in the higher-transmission regions to the transmission rate in the lower-transmission regions. The average transmission rate across all regions was kept fixed at 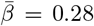 per day, corresponding to a basic reproduction number of 2. The simultaneous effects of heterogeneity and pandemic definitions in determining pandemic probability are illustrated in figure 6.

**Figure 6:**
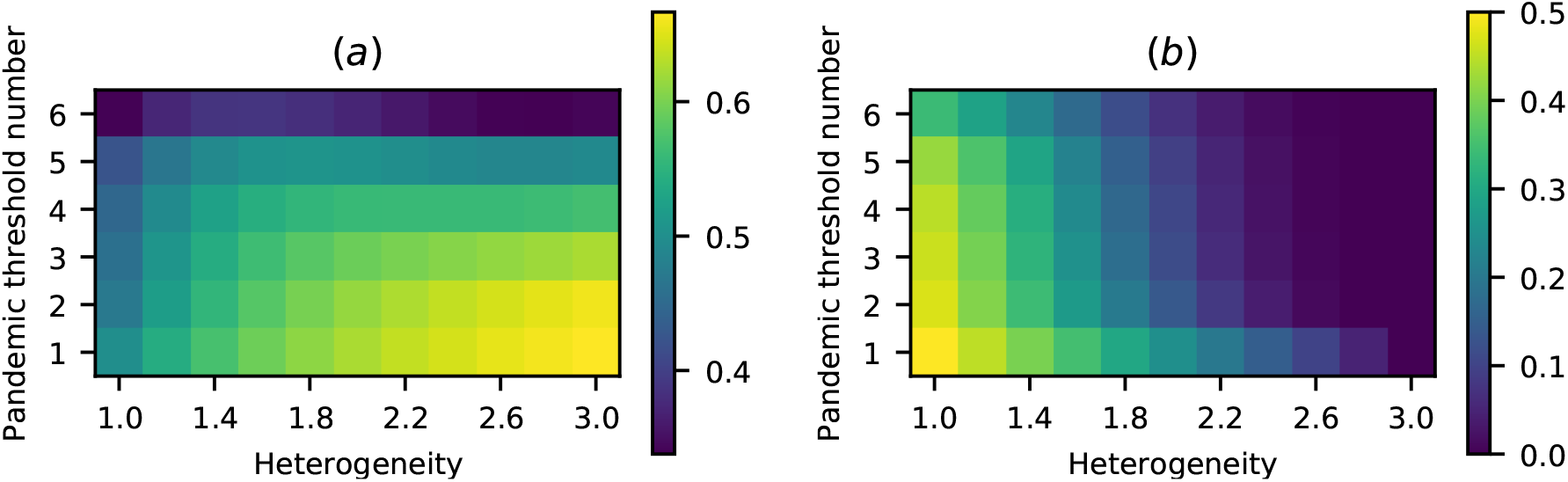
Pandemic probability for various degrees of heterogeneity of transmission rates and a range of pandemic definitions in terms of number of regions experiencing epidemics, on a fully connected six-region network where three regions are classed as higher-transmission and the other three regions are classed as lower-transmission. Note that the colour scales differ between the two plots, in order to make the variation in plot (a) clearer. a) Pandemic probability for a pathogen emerging in a higher-transmission region. For low thresholds heterogeneity increases pandemic probability, but at high thresholds pandemic probability grows and then decreases with increasing heterogeneity. b) Pandemic probability for a pathogen emerging in a lower-transmission region. At all thresholds increasing heterogeneity decreases pandemic probability.

The row for the 1-region definition shows how the risk of any outbreak varies with the changing basic reproduction number of the pathogen in the region in which it emerges. More complex effects can be seen for higher *n*-region definitions, especially five and six, where, at high levels of heterogeneity, even pathogens emerging in higher-transmission regions are prevented from spreading widely due to the low chance of epidemics in lower-transmission regions. Thus the probability of a pandemic under a 5-region or 6-region definition increases and then decreases with increasing heterogeneity. No corresponding effect exists for a pathogen emerging in a lower-transmission region, where increasing heterogeneity always decreases the chance of a pandemic, however it is defined.

## 3 Discussion

In this study, we have developed a theoretical framework to estimate the probability of a pandemic, as detailed in the Methods section below. We use a Markov chain technique based on SIR dynamics to model the spread of a pathogen. The results of this modelling framework reveal in which situations the definition of ‘pandemic’ has a strong effect on the calculated pandemic risk and in which situations it does not. The models also illustrate the effects of differing epidemiological parameters on pandemic risk under different definitions, and how these effects interact with each other.

Returning to the three epidemiological questions introduced in the introduction, we can see that our results show how the answers can depend on our definition of a pandemic, and on key population and pathogen parameters. The first question was “Would interventions restricting travel reduce the risk of a pandemic?” In figure 2, we see that reductions in travel rates always reduce risk in a sparsely connected network, where travel occurs mainly through a central hub. However, in a densely connected network with high travel rates, travel would have to be extremely highly suppressed to change the probability of a pandemic substantially, under most definitions. This accords with previous findings regarding the effectiveness of restricting travel [52]. Additionally, in the highly connected network, changing the definition of a pandemic makes little difference to the pandemic probability, for high enough values of the travel rate.

Figure 3 further illustrates the effects of different definitions. Changing the pandemic definition can sometimes greatly alter the estimated probability of a pandemic, as seen in figure 3a between the yellow line, representing the 2-region transregional definition, and the purple line, representing the 10-region transregional definition. The effect on the pandemic risk of reducing travel rates also differs greatly between to these two definitions. However, there are situations where changing the definition does not significantly change estimated pandemic probability, as seen in the same figure between the yellow line and the dashed green line, representing the 2-region interregional definition. In this case, both the estimated risk and the effect of reducing travel are very similar. So, while some changes in definition do not cause a large change in quantitative analyses of pandemic risk, others may significantly alter both our point estimates and the predicted effects of key parameters. Figure 3b shows that this may depend on the values of those key parameters themselves. For low travel rates, the two illustrated definitions diverge considerably, but at high travel rates the pandemic probabilities for the two definitions converge.

The second question was “Does a portion of the population have pre-existing immunity, and does this affect pandemic risk?” The presence of immunity can significantly alter the results discussed in the paragraphs above. In figure 5b, the leftmost column is equivalent to the column from figure 2b in which *λ* = 2.0 *×* 10^−4^ per day. However, as cross-immunity increases, a marked difference in the pandemic probability between different definitions becomes visible. This shows that the conclusion that precise pandemic definitions are of reduced importance in a highly connected network with high travel rates is context sensitive—if the population has high immunity, differences between definitions re-emerge.

The third question was “How is the risk of a pandemic affected by differences between regions?” In figure 6, we examined how heterogeneous transmission rates in different regions affect the risk of a pandemic. Many pathogens have higher transmission rates in lower income countries, and novel pathogens are more likely to emerge in low income countries [50, 51, 53]. Putting these two facts together, we see that pathogens are most likely to emerge in countries in which they have higher transmission rates. Motivated by this, we compared the scenarios of emergence in a higher-transmission and lower-transmission region, finding that pandemic definition makes a larger difference for diseases emerging in a higher-transmission region. In particular, when the pandemic definition requires many countries to experience epidemics to qualify an outbreak as pandemic, including countries with lower transmission rates, we see striking non-linearity in the relationship between pandemic probability and heterogeneity. For these definitions, as the difference in transmission rates between higher- and lower-transmission regions increases, the pandemic probability increases initially, before decreasing. This initial rise is due to the enhanced spread between high-transmission regions increasing the importation rate to low-transmission regions. This result implies that, when the mean value of the transmission rate is fixed, a small gap in the effectiveness of public health infrastructure between wealthy and poor regions puts all regions at greater risk, while a larger gap protects wealthier regions while the risk for poor regions continues to increase.

To illustrate this concept, consider the contrasting examples of Ebola and COVID-19. The 2014 outbreak of Ebola virus followed the pattern of high incidence in low income countries but low incidence in high income countries. It spread through several low-income African countries but was effectively contained when introduced to high-income countries [54–56]. In this case high-income countries had the capacity to prevent a pandemic from taking hold, being able to quickly isolate and treat symptomatic individuals. This generated high heterogeneity in transmission, corresponding to the right side of figure 6a, with low-income countries at high risk and high-income countries at low risk. In contrast, high-income countries have not been able to escape the pandemic of COVID-19, in part due to asymptomatic and presymptomatic transmission of SARS-CoV-2 allowing it to evade existing surveillance and public health measures [57, 58]. This has led to similar transmission rates across different countries, corresponding to the left side of figure 6a, where risk is more uniform between regions and therefore between pandemic definitions.

Further work could extend our modelling framework to investigate the role of pandemic definitions in quantifying the effects of additional epidemiological parameters on pandemic risk, such as use of different types of travel (e.g. within-country transport vs international flights) [45, 59, 60], the rate of nosocomial infections [61], or age-structured populations [62]. Our metapopulation modelling framework is generally applicable, and this framework could be extended to represent outbreaks of many different specific pathogens emerging in various locations.

In summary, we have developed a novel modelling framework for investigating pandemic risk. We have applied this framework to assess the pandemic risk in a range of different scenarios, and have interpreted the results under a variety of pandemic definitions. We have found that certain relationships, such as the effect of heterogeneity in transmission between regions on pandemic risk, are highly dependent on the definition of ‘pandemic’ used, while others, such as the effect of high travel rates on pandemic risk in a highly connected network, are not. This work provides a foundation for improved communication about pandemic risk, by highlighting the contexts in which pandemic definitions need to be provided in quantitative detail. In general, we contend that, when assessing the risk that an outbreak will develop into a pandemic, the precise pandemic definition used for a given analysis should be considered and stated clearly. Future work could investigate the effects of alternate definitions in more detailed epidemiological models, and extend this framework to investigate different dynamical features of pandemics.

## 4 Methods

We have combined standard epidemiological modelling techniques with a novel Markov chain treatment of metapopulation dynamics to produce a method of calculating the probabilities of epidemics and pandemics in a network of population regions. Suppose that we are treating the spread of a pathogen through *n* regions labelled *P*_1_, *P*_2_, *P*_3_, …, *P*_*n*_. Each region *P*_*j*_ has associated with it some intra-region pathogen transmissibility *β*_*j*_, disease recovery rate *µ*_*j*_, and population size *N*_*j*_. From these quantities it is possible to calculate a region-specific basic reproduction number *R*_0,*j*_. This can be fixed across all regions for a particular pathogen, or allowed to vary from region to region to reflect local epidemiological differences.

First let us consider the spread of the pathogen in a single region, using well-established results of stochastic Susceptible-Infected-Recovered (SIR) models. If a region *P*_*j*_ contains an initial number of infected individuals *I*_*j*_ (0), then in a discrete stochastic SIR model, the probability that these individuals do *not* cause an epidemic in *P*_*j*_ is 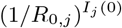 when *R*_0,*j*_ ≥ 1, and 1 otherwise. We also define the final size of an epidemic *R*_*j*_ (∞) (not to be confused with *R*_0,*j*_) as the number of recovered individuals in *P*_*j*_ at the end of the epidemic. This equals the total number of individuals in *P*_*j*_ who become infected at any time, and is given by the solution of the following equation [63].

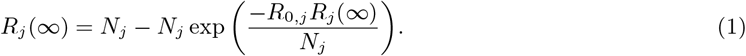

Now allow that infected individuals travel from region *P*_*j*_ to region *P*_*m*_ at a rate *λ*_*jm*_. We seek the probability that infected individuals travelling from *P*_*j*_ will *not* cause an epidemic in *P*_*m*_, in the case where initially infected individuals in *P*_*m*_ do not cause an epidemic in *P*_*m*_ (including the case where there are no initially infected individuals in *P*_*m*_). This is equal to the probability that *i* infected individuals migrate from *P*_*j*_ to *P*_*m*_, multiplied by the probability that this number of individuals fails to cause a major epidemic, summed over possible values of *i*. The minimum value of *i* is the case where no infected individuals migrate, and the maximum value is the case where all individuals in *P*_*j*_ that become infected at any point migrate. This gives us an expression for *q*_*jm*_, the conditional probability that, if *P*_*j*_ experiences an epidemic and *P*_*m*_ does not experience an epidemic due to a source of infected individuals other than *P*_*j*_, *P*_*m*_ does not experience an epidemic.

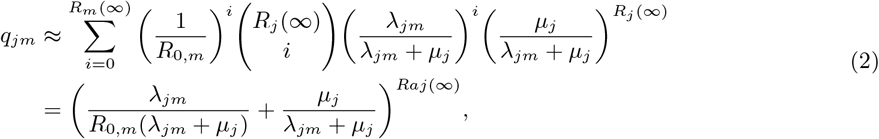

This approximation is valid when the number of infected individuals that travel between regions is much smaller than the size of the regions.

Let us assume that infected individuals travelling from a region *P*_*j*_ cannot cause an epidemic in a neighbouring region *P*_*m*_ if *P*_*j*_ does not itself experience an epidemic. Then computing the value of *q*_*jm*_ for every pair of populations *P*_*j*_ and *P*_*m*_ gives us sufficient information to determine the probability of any particular set of regions connected on a network experiencing epidemics so long as there are no interactions between different groups of migrants arriving in a region, and the total numbers of migrants in any region remains very small relative to the region’s size. If these assumptions hold, we can imagine the regions on a network with weighted directed edges, where the weight of the edge directed from region *P*_*j*_ to region *P*_*m*_ is *q*_*jm*_.

To determine how the final probabilities of epidemics depend on the pairwise probabilities *q*_*jm*_, we use a Markov chain. The states of this Markov chain assign one of three states to each region—*N* (for neutral), where it is not yet determined whether the region will experience an epidemic, *E* (for epidemic), where it is determined that the region will experience an epidemic but it is not yet determined in which further regions it will cause epidemics, and *T* (for terminal), where it is determined that the region will experience an epidemic and in which further regions it will cause epidemics due to onward transmission. As our model does not explicitly represent dynamical processes occurring over time, these states should not be interpreted as actual states of infection and recovery within regions, but rather as bookkeeping devices for the role of various regions in determining the spread of the pathogen through the network.

Suppose we have a network connecting *n* regions. In the initial state, each region where the initially infected individuals have caused an epidemic is in state *E*, and all the other regions are in state *N*. The global state of the network is simply the product of the states of each system. We can then define a transition matrix **T** that acts on the global state. The elements of this matrix are denoted 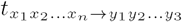.

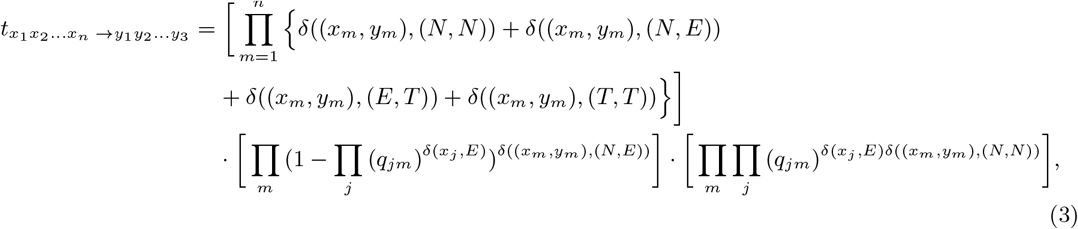

where

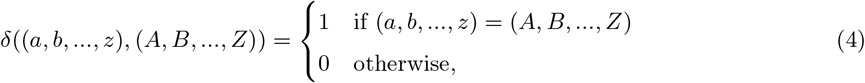

*x*_*j*_ is the state (*N, E*, or *T*) of region *P*_*j*_ before the transition, and *y*_*j*_ is its state afterwards. The expression inside the first set of square brackets ensures that the only acceptable transitions for any given region are *N* → *E* and *E* → *T*, and requires that all epidemic regions in the initial state must be terminated in the transition (this prevents double-counting of possible transmission paths). The expression inside the second set of square brackets gives the probability of each *N* → *E* transition, and the expression inside the final set of square brackets gives the probability of each *N* → *N* transition, given the regions that are in state *E* before the transition.

Note that these transitions do not represent a dynamical process—the order of transitions in this model does not necessarily correspond to the order in which regions experience epidemics. Instead, the transitions are simply stages along the exploration of different routes and outcomes from the disease spreading process.

The initial probability of each global state *z*_1_*z*_2_…*z*_*n*_ (where *z*_*i*_ ∈ {*N, E, T*}) is given by:

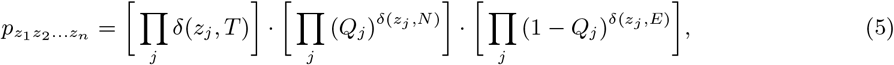

where 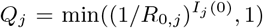 is the probability that the initial population of infective individuals does not cause an epidemic in region *P*_*j*_. Essentially, no region can begin in state *T*, and the probability of each initial global state is given by the product of the probabilities of each region being in the corresponding initial regional state.

In this system, all states in which no region is epidemic are absorbing, and in each transition at least one epidemic state must become terminal. This means that the system must reach an absorbing state in at most *n* transitions, since at least one region becomes terminal in each transition, and a fully terminal state is absorbing. So the final probability vector *p*_final_ is given by

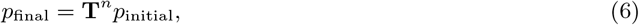

with **T** as the transition matrix and *p*_initial_ as the vector whose elements defined by equation (5). This final vector gives the probabilities of each configuration of the metapopulation, with populations in state *N* never experiencing an epidemic, and regions in state *T* experiencing an epidemic at some point.

### 4.1 Cross-Immunity

The model described above can incorporate certain epidemiological details, such as heterogeneity of population parameters, but is restricted to treating quite simple disease dynamics. In this section we expand the model to treat pathogens that give those who overcome infection cross-protection against future strains of that pathogen. This is necessary to be able to investigate how pre-existing immunity changes how pandemic definitions affect the results of our model.

We first describe the spread of a pathogen strain *X* using the methods above, introducing a superscript *X* to the relevant parameters to mark the strain, e.g. 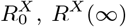, and 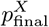. We assume that infection with pathogen *X* confers cross-immunity *α* to a second strain of the pathogen, which we call *Y*. In each population *P*_*j*_ we can define an effective basic reproductive number for *Y* in the case that *P*_*j*_ has experienced an epidemic of *X*, which we call 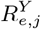.

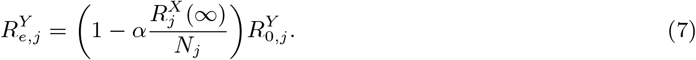

This expression simply multiplies the basic reproductive number by the effective number of susceptible individuals given the prevalence of cross-immunity in the population. It is through this expression that cross-immunity enters the model—the parameter *α* does not otherwise appear in what follows.

We can write down an equation for the expected total number of individuals in *P*_*j*_ infected in an epidemic of *Y* in analogy to equation (1). In the case where there has been no previous epidemic of *X* in *P*_*j*_, the expected epidemic size is the solution 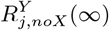 of

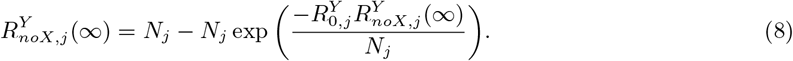

In the case where there has been a previous epidemic of *X* in *P*_*j*_, the expected epidemic size is the solution 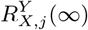 of

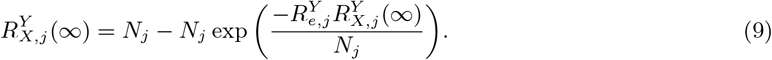

We assume that individuals infected with *Y* travel at the same rate as individuals infected with *X*. We then define the pairwise probabilities of transmission of *Y* between populations in analogy to equation (2). That is,

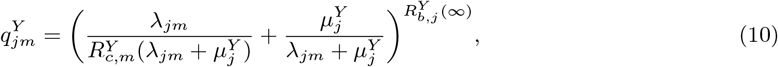

where 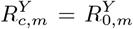 when *P*_*m*_ has not experienced a previous epidemic of 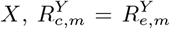 when *P*_*m*_ has experienced a previous epidemic of 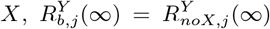 when *P*_*j*_ has not experienced a previous epidemic of *X*, and 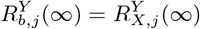 when *P*_*j*_ has experienced a previous epidemic of *X*.

These expressions for 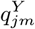 can be substituted for *q*_*jm*_ in equation (3) to yield a transition matrix for modelling the spread of *Y*, which we will call **T**^*Y*^ (*s*_1_*s*_2_…*s*_*n*_), where *s*_*j*_ is the final state (either *N* or *T*) of the *X* outbreak in *P*_*j*_. We find the initial probabilities of each state with regards to 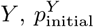, in analogy to equation (5), given an initial number of individuals infected with *Y* in each population 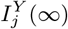.

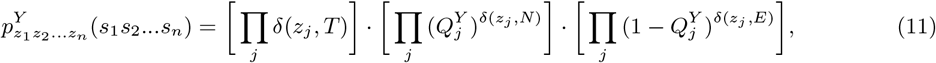

where 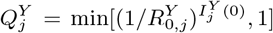 when *P*_*j*_ has not experienced a previous epidemic of *X* (i.e. *s*_*j*_ = *N*), and 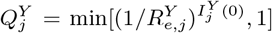 when *P*_*j*_ has experienced a previous epidemic of *X* (i.e *s*_*j*_ = *T*). We can then write the final probabilities of each combination of possible epidemics of *Y*, for a given set of previous epidemics of *X*, as

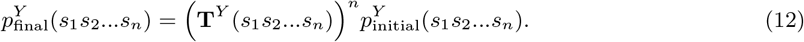

To find the overall probability of each combination of epidemics of *Y* in various populations given a prior probability of each combination of epidemics of *X* (given by 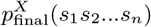 defined in equation (6)), we sum over the possible values of (*s*_1_*s*_2_…*s*_*n*_), weighted by their probability.

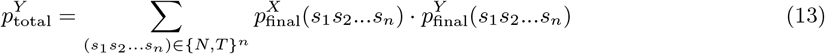

### 4.2 Code Availability

Code is available on the Open Science Framework at https://osf.io/z52te/.

## Data Availability

Code is available on the Open Science Framework at https://osf.io/z52te/.

## 5 Funding Acknowledgements

This work was supported by funding from the Biotechnology and Biological Sciences Research Council (BBSRC) [grant number BB/M011224/1]. This research was funded by Christ Church, Oxford, via a Junior Research Fellowship (RNT).

## 6 Author Contributions

BJS, RNT, and MBB conceived the study. BJS carried out the analysis, wrote the manuscript, and prepared the figures. All authors reviewed the manuscript.

## 7 Additional Information

The authors declare no competing interests.

## 8 Figure Legends

**1** Illustrations of a) a star network and b) a full network, each with ten regions. Circles represent regions, and straight lines represent travel routes between regions.

**2** Pandemic probability for a range of between-region transmission rates and a range of pandemic definitions in terms of number of regions experiencing epidemics. The “threshold number of regions” refers to the minimum number of regions that must experience epidemics before a pandemic is declared. Pandemic probability is in general sensitive to the pandemic definition used, but the degree of sensitivity depends on network structure and travel rates. a) Pandemic probability for a star network. Pandemic probability is in general highly sensitive to the pandemic definition used. b) Pandemic probability for a fully connected network. The sensitivity of pandemic probability to the pandemic definition used is significantly reduced at high travel rates.

**3** Plots of pandemic probability against travel rate for a range of pandemic definitions. The difference in probability for different pandemic definitions reduces as ravel rates increase. a) Plot of pandemic probability for a star network. b) Plot of pandemic probability for a full network. For a fully connected network all regions are adjacent, so no line is shown for non-adjacent regions.

**4** Plots of pandemic probability against travel rates for high and low values of cross-immunity on six-region networks. A pandemic is defined here as all six regions experiencing epidemics. The plots show a large relative difference both in likelihood of pandemics and in how that likelihood scales with travel rates. The initial infected individual for each outbreak appears in a randomly chosen region. a) Plot of pandemic probability for a star network. b) Plot of pandemic probability for a full network.

**5** Pandemic probability for various degrees of cross-immunity and a range of pandemic definitions in terms of number of regions experiencing epidemics, on a six-region network. a) Pandemic probability for a star network, assuming that the initial infected individual of the first epidemic is found in the central region, but allowing the second epidemic to emerge in any region with equal probability. b) Pandemic probability for a fully connected network. Here the sensitivity of pandemic probability to the pandemic definition used increases with cross-immunity, until the probabilty of any epidemic becomes very low.

**6** Pandemic probability for various degrees of heterogeneity of transmission rates and a range of pandemic definitions in terms of number of regions experiencing epidemics, on a fully connected six-region network where three regions are classed as high transmission and the other three regions are classed as low transmission. Note that the colour scales differ between the two plots, in order to make the variation in plot (a) clearer. a) Pandemic probability for a pathogen emerging in a high transmission region. For low thresholds heterogeneity increases pandemic probability, but at high thresholds pandemic probability grows and then decreases with increasing heterogeneity. b) Pandemic probability for a pathogen emerging in a low transmission region. At all thresholds increasing heterogeneity decreases pandemic probability.

